# Systematic review and network meta-analysis of drug combinations suggested by machine learning on genes and proteins, with the aim of improving the effectiveness of Ipilimumab in treating Melanoma

**DOI:** 10.1101/2023.05.13.23289940

**Authors:** Danial Safaei, Ali A. Kiaei, Mahnaz Boush, Sadegh Abadijou, Alireza Khorramabadi, Nader Salari, Masoud Mohammadi, Elham Parichehreh

**Affiliations:** Department of Electrical and Computer Engineering, University of Kashan, Kashan, Iran; Bioinformatics and Computational Biology (BCB) Lab, Sharif University of Technology; Cellular and Molecular Biology Research Center, Shahid Beheshti University of Medical Sciences, Tehran, Iran; Department of Computer Engineering, Shahid Bahonar University of Kerman, Iran; Department of Computer Engineering, University of lorestan, Khoramabad, Iran; Department of Biostatistics, School of Health, Kermanshah University of Medical Sciences, Kermanshah, Iran; School of Industrial Engineering, Iran University of Science & Technology

**Keywords:** Human genes, Machine Learning, Biological data analysis, Systematic Review, Network Meta-analysis, prescription drug information, Melanoma

## Abstract

**Background:** *<u>(Importance)</u>*One of the most dangerous kinds of skin cancer, Melanoma, develops in the cells (melanocytes) that make melanin, the pigment responsible for giving your skin its color. As well as developing everywhere on the body, including the eyes, Melanoma can sporadic occur internally, such as in the nose or throat. It is unknown what causes all melanomas, although exposure to ultraviolet (UV) radiation from the sun, tanning salons, and lamps increases the risk of getting them. As a result, radiation exposure increases the chance of obtaining Melanoma. Limiting your exposure to UV radiation can help reduce your risk of Melanoma. *<u>(Objective)</u>* Due to the unknown nature of this disease and its severe impact on human genes, the use of safe and effective drug combinations for treatment is very important. Proposed drug combinations should be administered with the greatest positive effect on the genes involved. Therefore, it is important to suggest an effective drug combination that can significantly affect the genes involved.

**Method:** *<u>(Data sources)</u>* This systematic review and network meta-analysis searched various databases, including Science Direct, Embase, Scopus, PubMed, Web of Science (ISI), and Google Scholar, without a lower time limit and up until July 2022, for articles focused on drug combinations for managing Melanoma. The study utilized a network meta-analysis to explore the effectiveness of the proposed medication combination on genes and proteins that may act as potential targets for improving Melanoma treatment.

**Results:** The results of this study show that the p-value between the proposed drug combination and Melanoma was 1.12E-08. This is while the p-value of Melanoma and only one drug has a maximum value of 0.0149. Therefore, the proposed drug combination’s effectiveness for treating Melanoma has increased 74 times. A systematic review has investigated the validity of the proposed drug combinations, human genes network meta-analysis, and prescription drug information.

**Conclusion:** The findings from this systematic review and meta-analysis suggest that the proposed drug combination reduces the p-value between Melanoma and genes that could potentially be targeted to slow the progression of the disease, ultimately improving its management. Therefore, selecting the appropriate drug combination is critical for enhancing community health and reducing per capita treatment expenses.

**Highlights:**

- Melanoma is one of the most aggressive kinds of skin cancer, and it begins in the cells (melanocytes) responsible for producing melanin.
- Therapy must make use of pharmacological combinations that are both safe and effective.
- Any proposed medication combinations must be delivered in a way that will have the maximum possible beneficial impact on the genes at play.
- In this research, an effective pharmacological combination for the treatment of melanoma illness is investigated.
- The results suggest that the suggested treatment combination is beneficial in the treatment of Melanoma, as it reduces the p-value between the disease and the genes identified as potential targets for therapy. This indicates that the proposed treatment approach has the potential to improve the management of Melanoma.

## INTRODUCTION

This study comprised four key stages: extracting drug combinations through the use of native machine learning, conducting a systematic review, performing a network meta-analysis on human genes, and gathering information on prescription drugs (Kiaei et al., 2022). The research followed a four-stage process, beginning with the use of a native machine learning system to suggest a drug combination for treating Melanoma. Next, the drugs proposed in this combination underwent a systematic review. The third step involved a network meta-analysis to assess the efficacy of this proposed drug combination on human genes (Salari et al., 2021). The fourth and final stage involved examining the prescription drug information for the identified drug combinations, which included evaluating factors such as drug interactions, potential side effects, and compatibility with other drugs and foods.

The study conducted a systematic review to evaluate the soundness of the suggested drug combinations, the network meta-analysis conducted on human genes, and the prescription drug information.

Malignant Melanoma, often known as Melanoma, is a kind of cancer that inspirational motivation in the skin and affects body areas that have skin. It might begin in a mole or in the skin that appears normal. Melanoma is not a recent illness; nonetheless, there is little historical documentation of its prevalence. One example is the investigation of nine Peruvian mummies in the 1960s. These mummies were radiocarbon dated to be around 2,400 years old, and they all showed obvious melanoma symptoms, including melanotic tumors in the skins and dispersed metastases to the bones. There are several causes of Melanoma. Radiation exposure can be listed as one of them. Melanoma often affects the skin, but it can also affect other regions of the body. For instance, it sometimes affects the mouth, intestines, and eyes (uveal Melanoma). It appears in various locations for both men and women. Women often have them on the legs, but men typically have them on the back. Moles are responsible for around 25% of melanomas, and skin alterations in moles will spread to other body parts. An increase in size, uneven margins, discoloration, itching, or skin disintegration in a mole are changes that could indicate Melanoma.

### Rationale

Several drugs have been suggested to block the receptors of these human genes and treat Melanoma. These drugs include Ipilimumab, InterferonAlfa-N1, Trametinib, Sargramostim, and Vemurafenib, with Table 1 providing information on some of their properties from Drugbank. Some of these drugs have been identified in multiple articles as effective treatments for Melanoma, including:

**Table 1:**
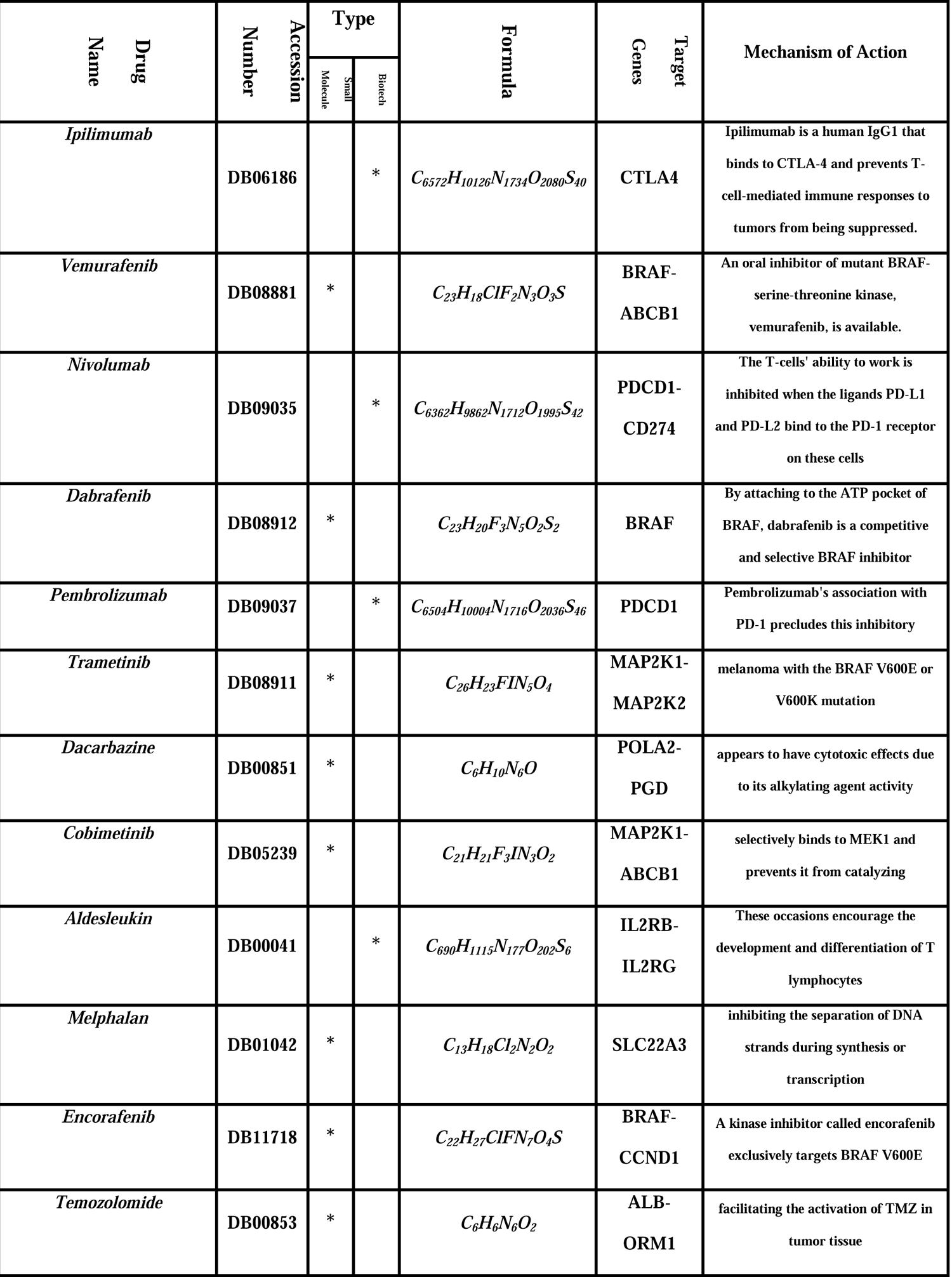
properties of some drugs that are mentioned in many studies in melanoma treatment.

*Ipilimumab* is a human cytotoxic T-lymphocyte antigen 4 (CTLA-4) blocking antibody used to treat metastatic or unresectable Melanoma.

*Interferon Alfa-N1* is a purified form of human interferon used to stimulate the innate antiviral response in treating genital warts due to the human papillomavirus.

*Trametinib* is a kinase inhibitor used to treat patients with specific types of Melanoma, non-small cell lung cancer, and thyroid cancer.

*Sargramostim* is a modified form of recombinant human granulocyte-macrophage colony-stimulating factor used to increase immune cell production after myelosuppressive therapy or bone marrow transplant.

*Vemurafenib* is a kinase inhibitor used to treat patients with Erdheim-Chester Disease who have the BRAF V600 mutation and Melanoma in patients with the BRAF V600E mutation.

Each of the drugs listed only affects a limited number of human genes linked to Melanoma, underscoring the importance of identifying the appropriate combination of drugs that can target a greater number of genes than those listed above (Salari et al., 2023b). The study employed a native machine learning model to investigate drug-based treatment strategies for combating Melanoma.

### Objectives

Considering several reported Melanoma treatment consequences of drugs (Figure 1), In order to deliver a comprehensive and meticulous network meta-analysis of genes/proteins and their response to drug combinations for Melanoma treatment, a systematic review of previous research in this area was conducted. The study was driven by the absence of worldwide data and general statistics on the subject matter. The objective of this study is to extensively investigate, evaluate, and analyze published studies on the impact of suggested drug combinations (Salari et al., 2022b) on the management of Melanoma.

**Figure 1:**
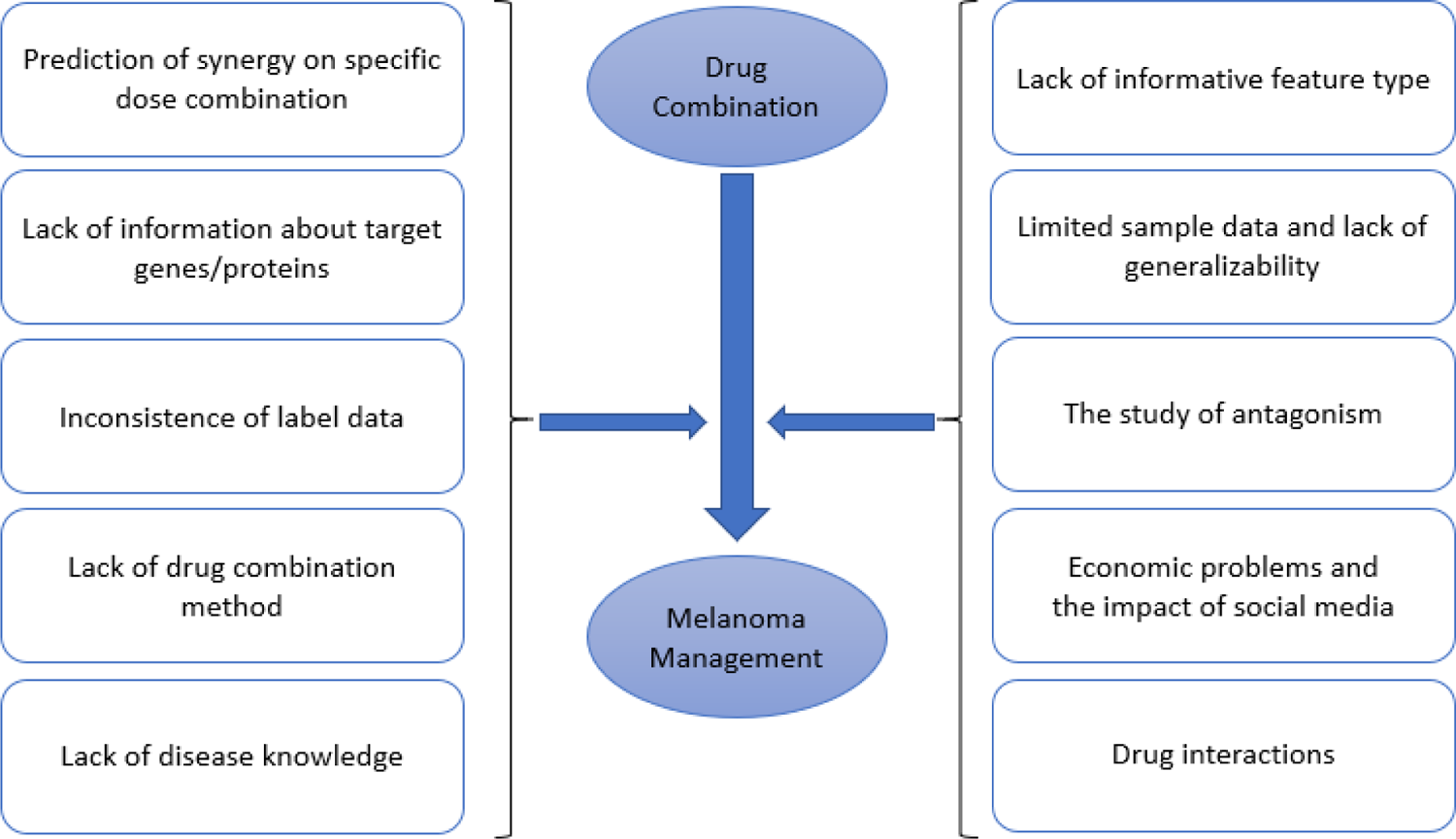
Impacts of suggested drug combinations on Melanoma treatment

## METHODS

### Information sources

To validate the proposed drug combination generated by our native machine learning analysis model, we conducted a systematic review using NLP techniques to identify relevant research in Science Direct, Embase, Scopus, PubMed, Web of Science (ISI), and Google Scholar databases. Although the machine learning model provided the suggested drug combination, the focus of our study was on verifying its effectiveness. To do this, we extracted keywords from the Melanoma subscription and the outputs of the machine learning model from the first step, which included Melanoma, Ipilimumab, Interferon Alfa-N1, Trametinib, Sargramostim, and Vemurafenib.

### Search strategy

The NLP technique used in this study searches for relevant articles by semantically analyzing their Titles/Abstracts and connecting to databases such as Science Direct, Embase, Scopus, PubMed, Web of Science (ISI), and Google Scholar. MeSH terms are also considered during the keyword search. For instance, the term Melanoma is equivalent to several related terms such as malignant melanoma, melanocarcinoma, and nevocarcinoma. There is no restriction on the timeframe for searching the relevant articles.

### Eligibility criteria and Selection process

It seems like you have provided a clear description of the inclusion and exclusion criteria for selecting studies for your systematic review. Including only studies that meet these criteria can help ensure the quality and relevance of the evidence used to support your analysis. By excluding interventional studies and focusing on observational research, you can also limit the potential for bias that may arise from experimental interventions or the influence of treatment on study outcomes. This approach can enhance the validity of your findings and provide a more accurate representation of the effects of the proposed drug combinations on Melanoma.

### Study selection

The systematic review process involves filtering out redundant studies and evaluating the remaining studies in a step-by-step manner using a list of their names. The title and abstract of each study are carefully examined in the screening stage, and studies that do not meet the selection criteria are excluded. Then, the full text of the remaining studies is thoroughly analyzed in the competency evaluation stage to eliminate irrelevant studies. To prevent personal bias in the selection process, an expert and an NLP Question-Answering agent independently conduct research and data extraction. The expert provides accurate reasons for excluding studies that do not meet the criteria. In addition, a QA agent assesses each article based on specific questions and assigns a score. Articles that receive low scores will be removed from consideration. The questions posed by the QA agent focus on the effectiveness of each drug in managing Melanoma, and each drug suggested by the ML model is evaluated individually. If there is any disagreement between the expert and the QA agent’s output, the expert will review the relevant article. After these steps, 29 studies were selected for the third stage of the systematic review.

### Quality evaluation

To assess the quality of the selected papers based on their research type, a checklist was developed that evaluated both methodological validity and findings. Typically, STROBE checklists are utilized to assess the efficacy of observational studies. The checklist consisted of six general scales/sections, including the title, abstract, introduction, methodology, results, and discussion, with some scales having sub-scales, resulting in a total of 32 fields (sub-scales). Each of these 32 areas represented distinct components of a research methodology, such as the title, problem statement, study goals, type of study, statistical population, sampling method, sample size, description of variables and procedures, data collection method(s), statistical analysis methods, and conclusions.

The use of the STROBE checklist for qualitative assessment during the research phase resulted in a maximum score of 32. Articles with a score of 16 or above were considered of moderate to high quality, while those scoring below 16 were categorized as poor quality and excluded from the study. Out of the initial pool, 29 articles met the criteria for moderate to high quality and were included in the systematic review. This is in line with the data collected in the previous steps. The research protocol for this article is based on RAIN (Salari et al., 2023b). Numerous research studies have employed the RAIN protocol as a framework for their research(Boush et al., 2023; Jafari et al., 2022a; Kiaei et al., 2022, 2023, n.d.; Salari et al., 2021, 2022b, 2022b, 2023a).

### Study risk of bias assessment

At this stage, the effectiveness of each drug on human genes is calculated based on the p-value. The p-value is used to determine the efficacy of each medication on human genes at this point. Circular bar charts, radar charts, and Table 5 will display the findings.

## RESULTS

This investigation examined how the mentioned drugs impacted the treatment of Melanoma. Following the PRISMA criteria, relevant research papers on this subject were systematically gathered with no time limit up to July 2022. From the initial search, 329 articles were identified and stored in the reference management tool EndNote. Of these, 198 were duplicates and removed. The remaining 131 papers were screened based on their titles and abstracts, and 28 were eliminated. During the eligibility evaluation stage, 67 of the 103 remaining articles were excluded after analyzing their entire texts and considering the inclusion and exclusion criteria. At the quality assessment phase, 29 cross-sectional studies were chosen after four studies with poor methodological quality were excluded based on their STROBE checklist scores. (Please refer to Figure 2). The details and characteristics of these studies are also presented in Table 2.

**Figure 2:**
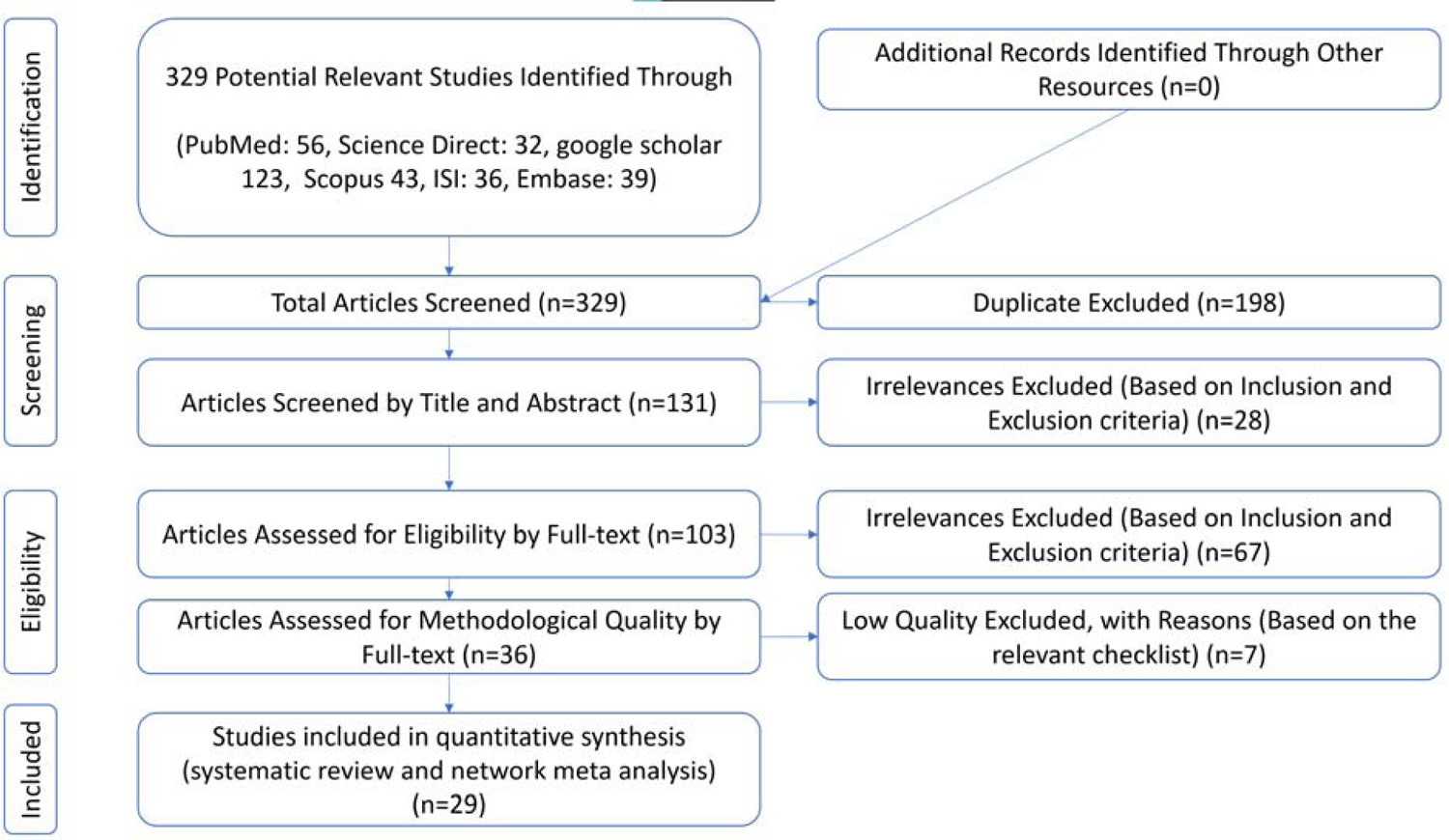
The phases of screening articles in this systematic review and network meta-analysis are shown in PRISMA’s (2009) flow diagram.

**Table 2:**
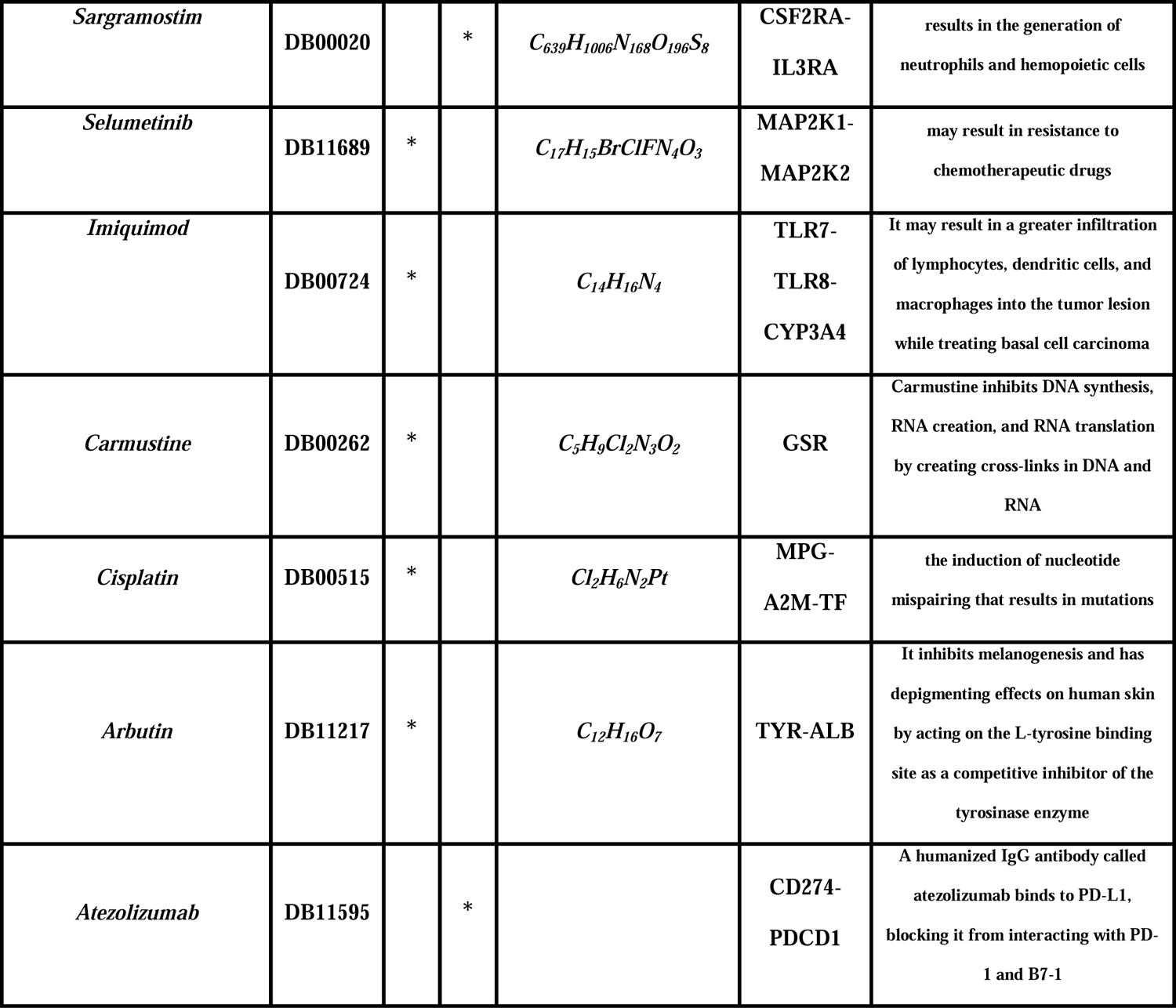
some important research studies for proposed drugs in Melanoma treatments

### Genes Network Meta-Analysis

Table 3 displays the p-values resulting from combining the drugs mentioned above. For example, Scenario 2, which combines Melanoma with Ipilimumab and Interferon Alfa-N1, had a p-value of 0.000602, which decreased to 0.000029 when Trametinib was added to the combination in Scenario 3. Additionally, Table 3 shows that the proposed drug combination had a positive effect on treating the disease, as demonstrated by the p-value after applying the fifth scenario.

**Table 3:**
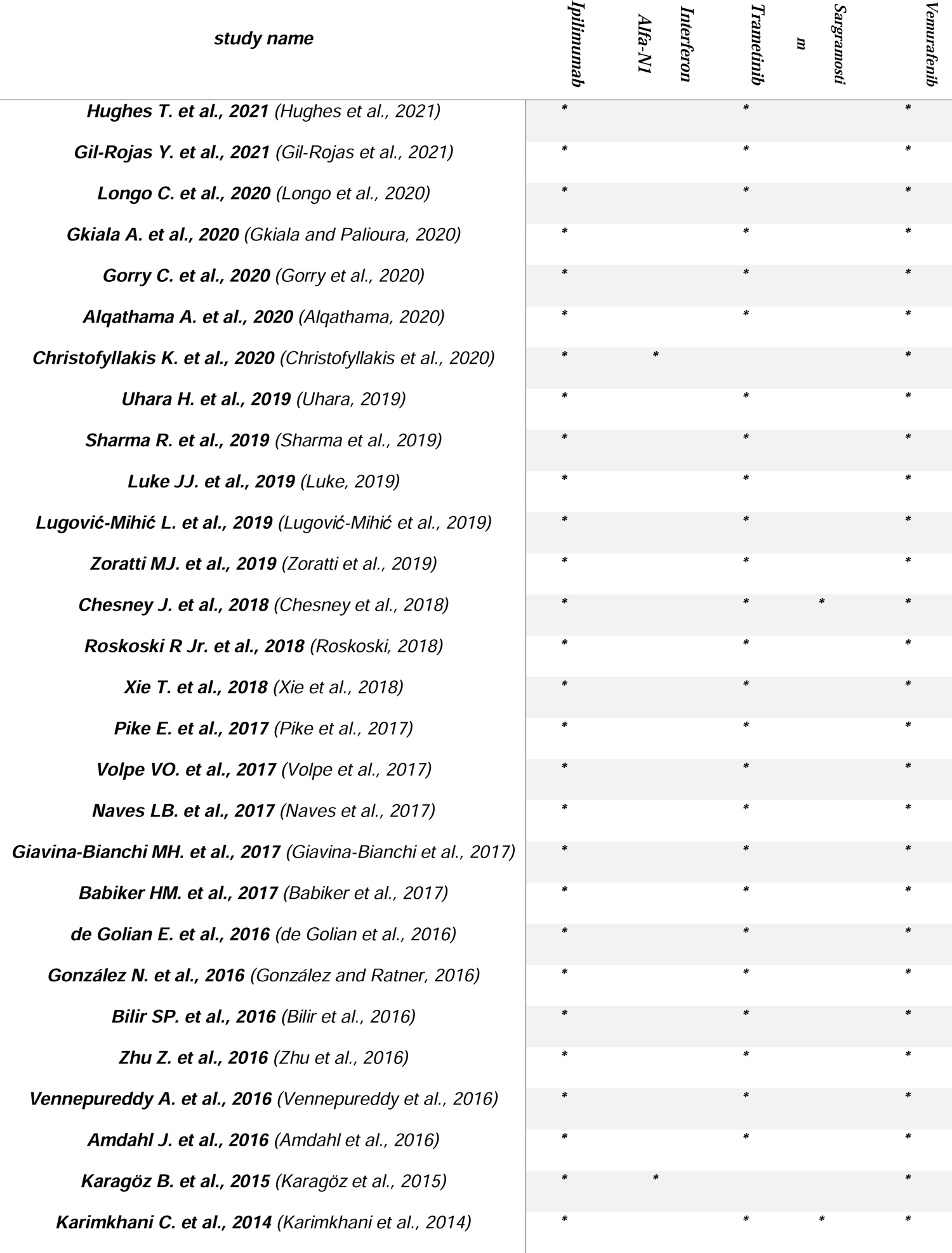
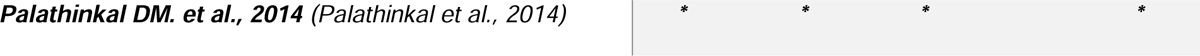
p-value between scenarios and Melanoma

Table 4 demonstrates the changes in p-values between human genes and Melanoma resulting from new scenarios. The “S0” column shows the p-value between Melanoma and the corresponding affected human genes. The “S1” column displays the combined p-value between Melanoma and the human genes where Ipilimumab is utilized. In the “S5” column, which includes the scenarios defined in Table 3, the p-values between Melanoma and many human genes reach 1.

**Table 4:**
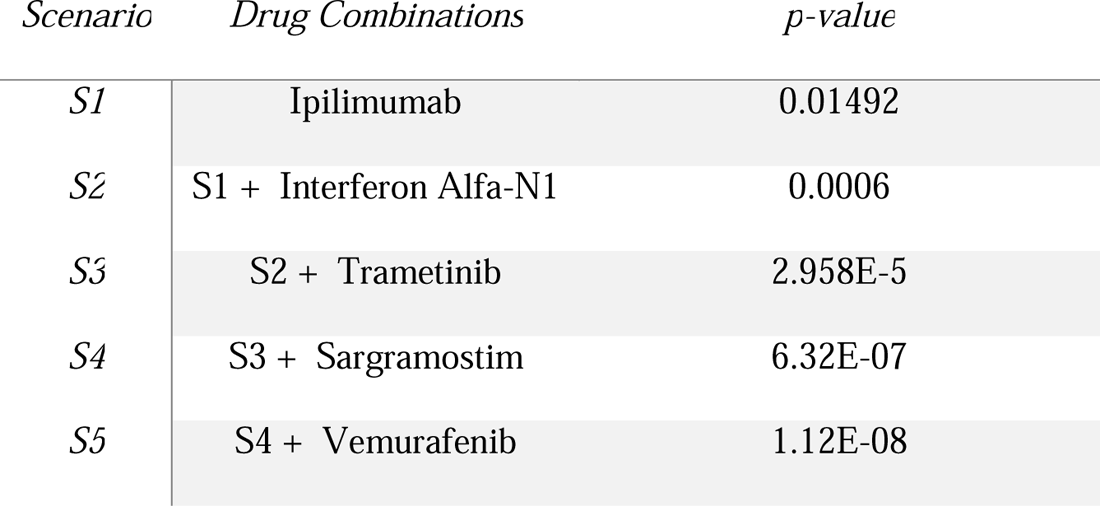
p-values between Melanoma and human genes before/after different scenarios

**Table 5:**
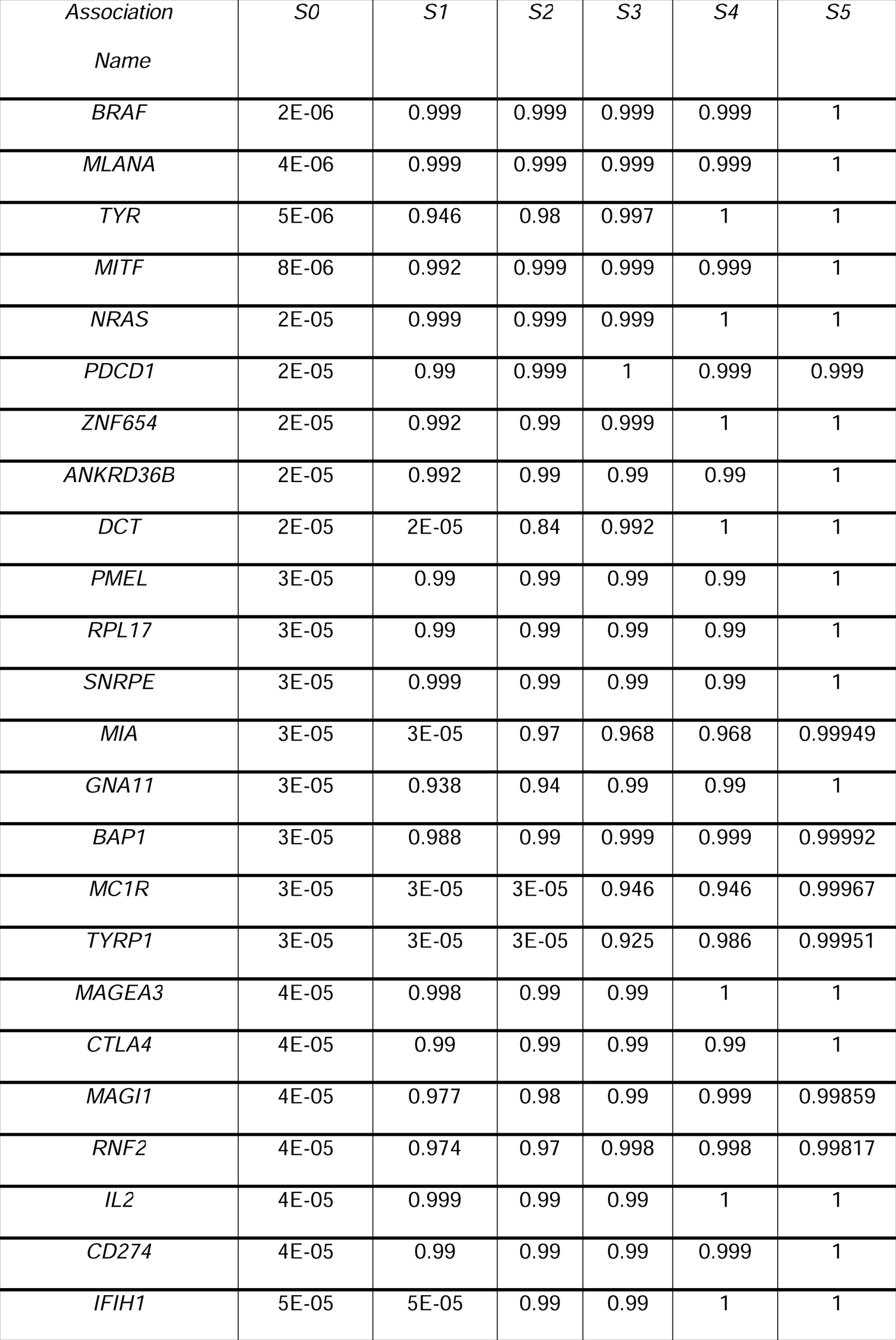
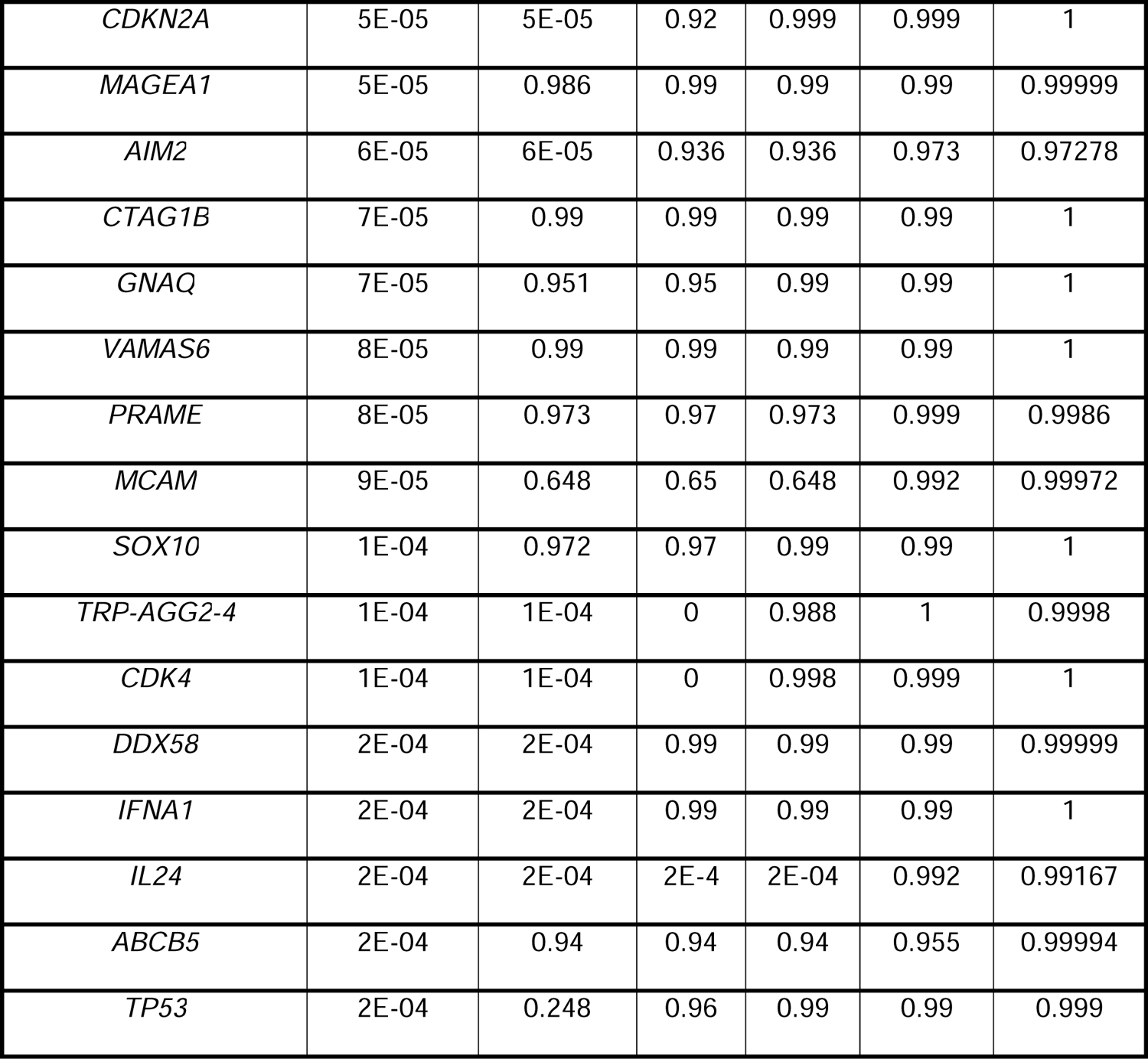
properties of proposed drugs from drugbank

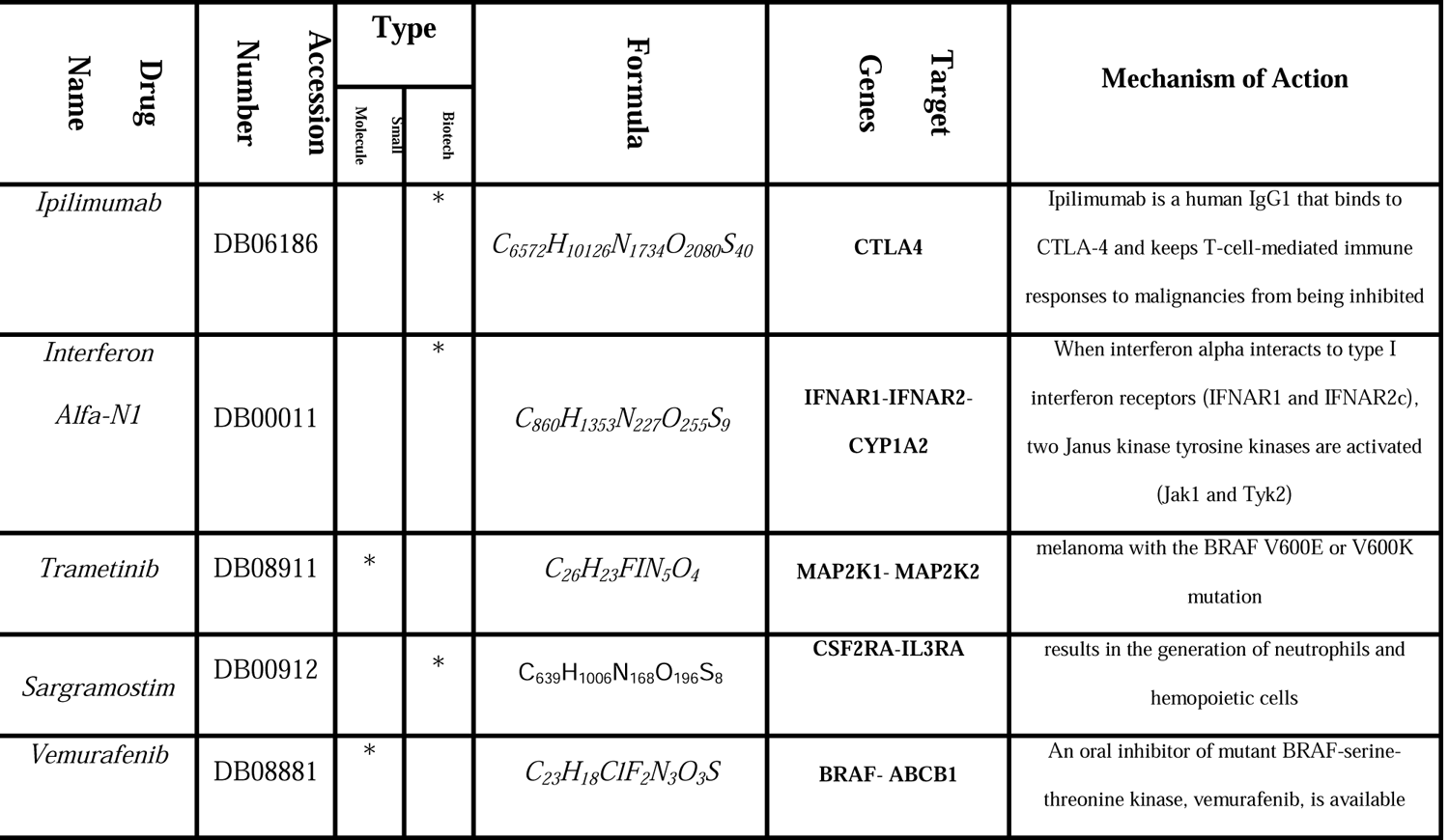

Figure 3 (a) depicts the p-values between affected human genes and Melanoma, while Figure 3 (b) illustrates the p-values between them after implementing the fifth scenario. Additionally, Figure 4 uses a radar chart to show the efficiency of the drug selection algorithm by presenting the p-values between Melanoma and human genes after the selected drugs have been administered. Each colored line represents the effectiveness of the corresponding drug in that particular scenario.

**Figure 3:**
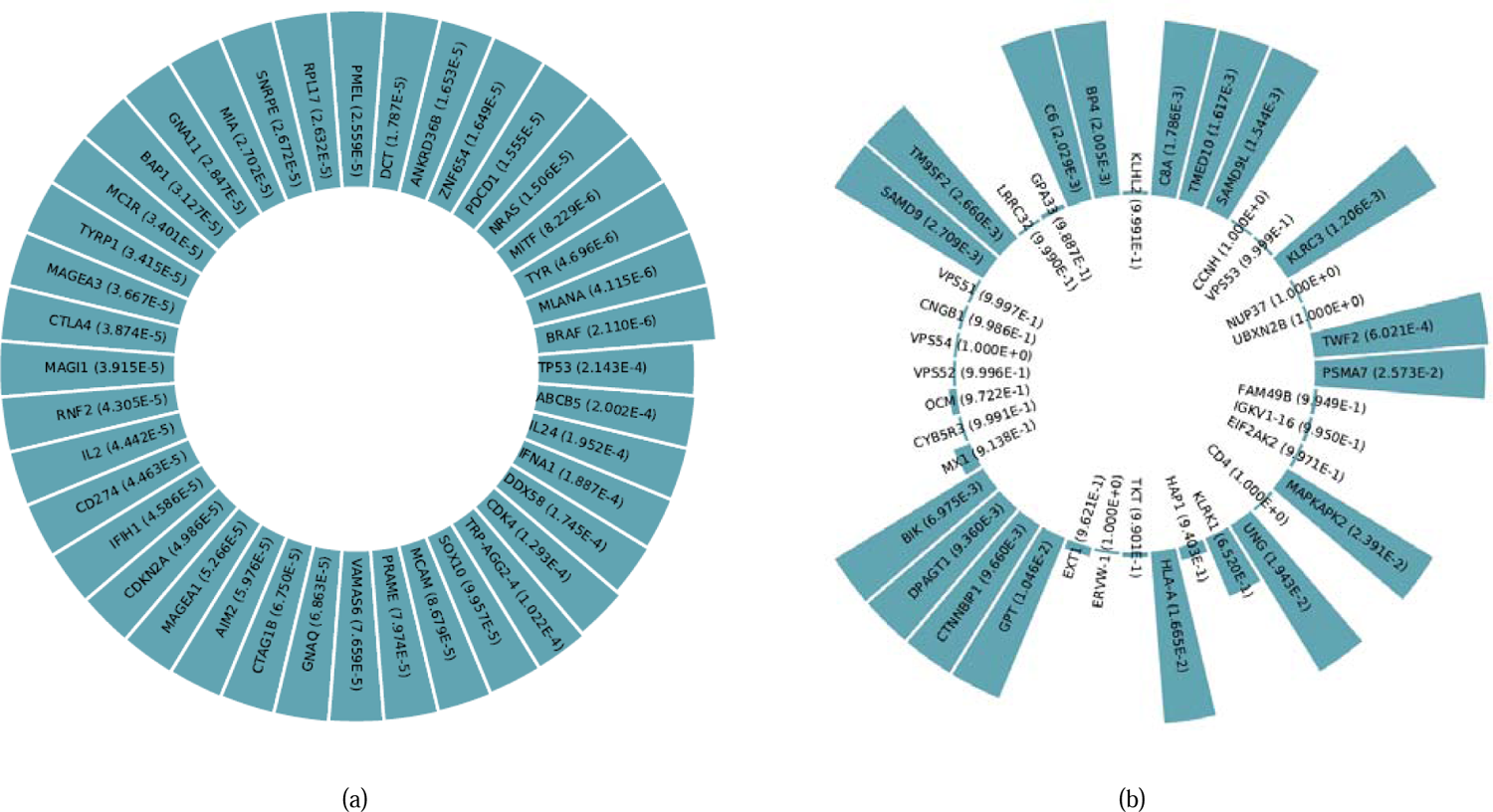
p-values between affected human genes and Melanoma {(a)before/ (b)after} implementing Scenario 5.

**Figure 4:**
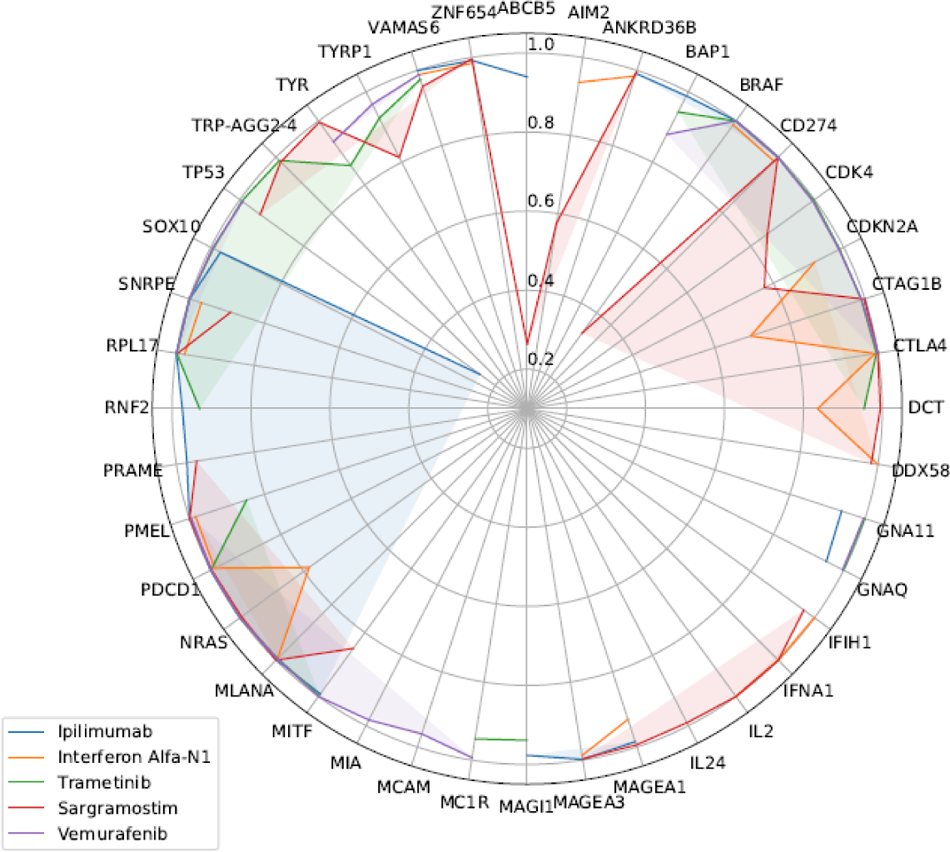
radar chart for p-values between Melanoma and affected genes, after consumption of each drug.

## DISCUSSION

The cells that make melanin, the pigment that gives your skin its color (melanocytes), which is the most dangerous kind of skin cancer, are where Melanoma begins. Additionally, Melanoma can develop in your eyes and, very rarely, within your body, like in your throat or nose.

The actual reason all melanomas occur is unknown, although exposure to ultraviolet (UV) radiation from sunshine, tanning beds, or lamps increases your chance of getting the disease. You can lower your melanoma risk by limiting your exposure to UV light. Paleopathologists in Peru examined mummies from the fourth-century bc and discovered spherical melanotic tumors in the skin and widespread metastases in the extremity and skull bones. Hippocrates (460–375 bc), then the Greek physician Rufus of Ephesus (60–120 ad), provided the earliest recognized description of Malignant Melanoma. Other medical professionals have documented pigmented skin cancers with distant metastases throughout history. However, important advancements in recognizing and controlling human malignant Melanoma did not occur until the 1800s. Laennec examined “la melanosis” and referred to its hue as melanotic in an unpublished report he sent to the Faculté de Médecine in Paris in 1806. Norris provided the first explanation of the genetic causes of Melanoma in 1820. 5 In 1838, Carswell used the word “melanoma” to refer to pigmented malignant skin tumors. 6 Pemberton promoted and carried out extensive and deep radical excision for Melanoma in 1858. This idea, which emerged a full century before it was adopted as a surgical method, also promoted groin dissection to remove all affected lymph nodes. Handley once more promoted the en bloc excision of Melanoma with broad margins in 1907, according to the dissection in continuity approach. One instance, however, is from a 1960s investigation of nine Peruvian mummies that were radiocarbon dated to be around 2400 years old and displayed apparent melanoma symptoms, including melanotic tumors in the skin and widespread metastases to the bones. This study employed a machine learning-based system to identify effective drug compounds for treating Melanoma, with the aim of enhancing the treatment process.

The use of safe and effective drug combinations is crucial for treating Melanoma, given its unknown nature and the potential impact on human genes. In this study, a native machine learning model was utilized to propose effective drug combinations. The research consisted of four main steps: drug combination extraction using the machine learning model, systematic review, human genes network meta-analysis, and prescription drug information analysis.

The proposed drug combinations were validated through a systematic review, genes network meta-analysis, and investigation of prescription drug information. The study found that the proposed drug combinations covered a wider range of human genes involved in Melanoma and were effective in treating the disease.

### Related genes/proteins

The relevance of genes in Melanoma progression has been extensively studied and validated in numerous studies. These studies have highlighted the involvement of various genes and proteins in the disease. Given that the focus of the article is on validating the proposed drug combination rather than the machine learning model, a systematic review was employed to verify the results. Systematic reviews are commonly used in research to validate claims regarding the relationship between two or more subjects. (Salari et al., 2021) Demonstrates the incidence of osteoporosis among older persons worldwide using a thorough systematic review and meta-analysis. (Salari et al., 2022a) Research studies that employ systematic review methodology to investigate Duchenne and Becker muscular dystrophy are widely conducted around the world.

### Details of the Method

The initial stage of this study involved the use of a native Machine Learning (ML) model (Jafari et al., 2022b) to propose various drug combinations for the treatment of Melanoma. The ML model takes in p-value information regarding the relationship between the disease and the relevant biological data, specifically human genes, as well as the p-values between the same human genes and effective drugs as input. It then outputs a suggested drug combination that will drive the p-value between the disease and the same human genes closer to 1.

Using NLP-based semantic search methods to extract articles for the systematic review can save a considerable amount of time and ensure a wider range of articles are considered. MeSH terms are an effective way to capture relevant articles and can help to reduce the likelihood of missing important studies. This approach also allows for more comprehensive and accurate searches, as it includes related keywords that may not have been considered in a manual search. However, it is important to note that this approach still requires careful review and screening of articles to ensure they meet the inclusion criteria for the study.

In the fourth step, prescription drug information for the proposed drug combinations is investigated. This includes drug interactions, side effects, contraindications, drug-food interactions, and other relevant information. This information is important to ensure the safety and efficacy of the proposed drug combinations. The investigation is done through various sources such as drug databases, clinical studies, and package inserts. The information gathered in this step is then analyzed to ensure that the proposed drug combinations are safe and effective for use in treating Melanoma. Overall, the four-step approach in this study provides a comprehensive and systematic method for identifying and validating effective drug combinations for Melanoma treatment.

### Proposed drug combination using our ML model

Machine learning has been utilized in numerous medical studies. One instance is the utilization of deep learning and Shapley values in (Jafari et al., 2022b), where a comprehensive diagnostic and predictive pipeline for identifying the risk of chronic diseases in cohort research was developed. In this phase, the native ML model obtains a weight between Target (Melanoma) and Associations (Drugs) as input through an Interface Feature, which is biological data (human gene). The p-value is used to calculate this weight. Figure 5 illustrates the architecture of our native ML model, which begins by considering the p-value between the Target and human genes. For example, p^FT^ Represents the p-value between the human gene F1 (as interface feature) and the MELANOMA (as Target). Additionally, this model also receives the p-value between the same human genes and different drugs. (For example, p ^AF^ represents the p-value between the first association and the third Interface human gene.) Using the above biological data as inputs, the ML model estimates the combined p-value between Associations and the Target. (For example, cp^AT^ represents the combined p-value between the second Association and Target.)

**Figure 5:**
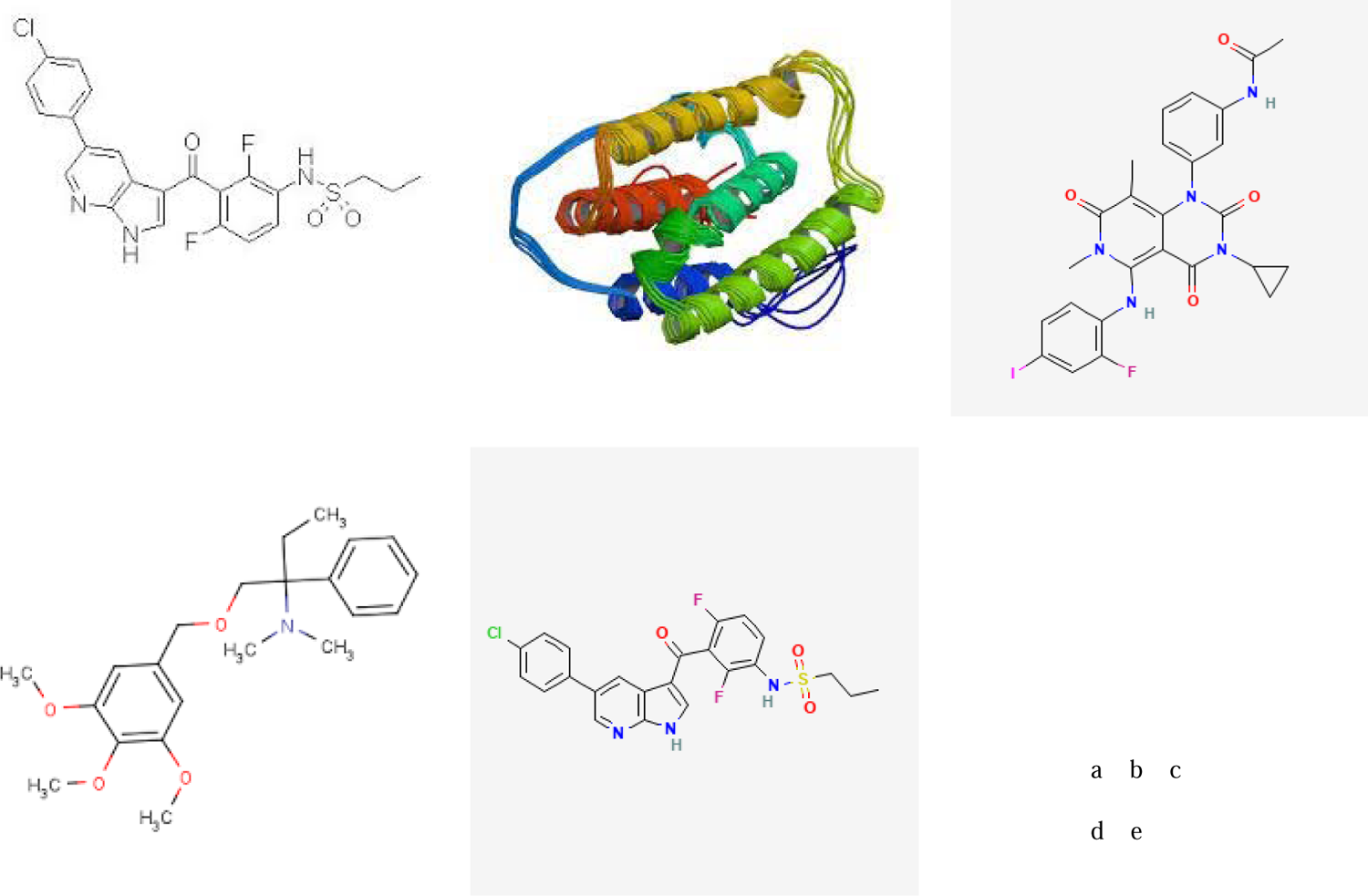
Drug structure for (a) Ipilimumab, (b) Interferon Alfa-N1, (c) Trametinib, (d) Sargramostim, (e) Vemurafenib from https://www.drugbank.com/

**Figure 6:**
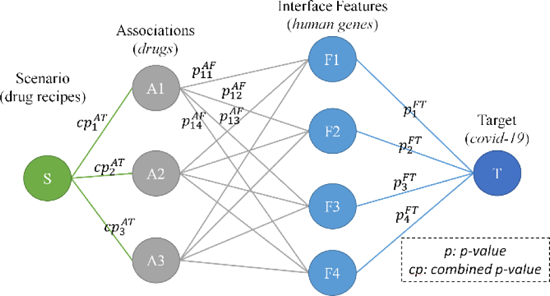
The general structure of the native ML model to suggest an effective drug combination in the management of disease using human genes as interface features

The ML model then selects the association with the lowest combined p-value and updates all weights based on its effect on the interface features. The goal is to make other associations have a similar effect on the human genes as the selected association and increase their p-values. This iterative process is repeated until the ML model reaches its stop condition.

Once the first drug is selected, the algorithm updates the weights between MELANOMA and affected human genes based on the p-values between those genes and the selected drug. This process is repeated, and the algorithm selects the next drug with the lowest combined p-value. The weights are then updated again, and the process continues until the algorithm reaches its stop condition. By selecting drugs with low p-values, the ML model identifies drug combinations that have the potential to effectively treat MELANOMA.

As a result, the drug selection algorithm updates the combined p-values between the selected drugs and MELANOMA, and repeats the process of finding the drug with the lowest combined p-value and updating weights.

Ultimately, the algorithm generates multiple scenarios, each containing a drug combination, which is the main objective of the algorithm in facilitating cooperative associations.

### Systematic Review

However, the drug combinations are provided by the native machine learning approach; current work innovates a particular presentation of the drug combinations. Hence, a systematic review is employed to validate the study. As shown in Table 2, the indicated articles have been investigated to confirm subgroups of the proposed drug combination.

### Prescription medication details

Certain prescription drug information is utilized to examine interactions between drugs, drug-food interactions, potential side effects, and the risk of serious conditions.

Reputable websites such as Medscape^8^, WebMD^9^, Drugs^10^, and Drugbank, ^11^ have conducted investigations into drug interactions, testing the drugs in pairs.

The internet databases, including Medscape, WebMD, Drugs, and Drugbank, analyzed the drugs in pairs to identify potential drug interactions. Based on the information retrieved from these databases, several drug interactions were identified among the mentioned drugs, which will be discussed below.

### Drug interactions

Between Ipilimumab and Vemurafenib, we will have a moderate drug interaction, which has been investigated. Ipilimumab may increase the hepatic activities investigated. Ipilimumab use of vemurafenib and Ipilimumab may increase the incidence and/or severity of hepatotoxicity, including increased liver transaminases and bilirubin. While prescribing information for vemurafenib cautions against concurrent use of vemurafenib and Ipilimumab, further studies have shown that the benefits of combination therapy may outweigh the risks.(Amin et al., 2016; Bunchorntavakul and Reddy, 2017; Ribas et al., 2013)

Minor medication interactions with Ipilimumab and trametinib have also been documented. Trametinib elimination could be slowed down by Ipilimumab, which could result in higher serum levels. Several kidney functions work together to produce renal excretion of medications, including glomerular filtration, passive diffusion, tubular secretion, and tubular reabsorption. Therefore, They are vulnerable to competition from various substrates that the kidneys discharge. The kidneys’ largely excreted drugs may compete with one another if given simultaneously.(Popović et al., 2016; Tiong et al., 2014; van Ginneken and Russel, 1989) One drug will likely “out-compete” or saturate the renal excretion mechanisms before eliminating other drugs. Consequently, eliminating these other co-administered drugs may be inhibited or delayed, leading to increased serum concentrations and the risk, incidence, and/or severity of adverse effects associated with exposure to these drugs. (Tiong et al., 2014)

## CONCLUSION

This study proposed a drug combination consisting of Ipilimumab, InterferonAlfa-N1, Trametinib, Sargramostim, and Vemurafenib for treating Melanoma. The p-value between Melanoma and the human genes associated with this combination has reached 0.015, which indicates that this drug combination is very effective. Other antiviral therapies are still debatable regarding their efficacy and safety, and further high-quality clinical trials are needed.

## Abbreviations

STROBE: PRISMA: Preferred Reporting Items for Systematic Reviews and Meta-Analysis; Strengthening the Reporting of Observational Studies in Epidemiology

RAIN: Systematic Review and Artificial Intelligence Network Meta-Analysis

## Data Availability

Not applicable

## Acknowledgments

This research is performed by Student researchers in Bioinformatics, Computational Biology, Machine Learning, and Medicine.

## Authors’ contributions

NS contributed to the design, AAK, MB and DS native machine learning algorithm, MM, AAK, MB, and DS biostatistical analysis; SA and DA took part in most research procedures. SA, DA and AK utilized machine learning to investigate medication combinations. The manuscript’s content has been viewed by all writers and is now their opinion.

## Availability of data and materials

Datasets are available through the corresponding author upon reasonable request.

## Ethics approval and consent to participate

Not applicable.

## Consent for publication

Not applicable.

## Footnotes

8 https://reference.medscape.com/drug-interactionchecker

9 https://www.webmd.com/interaction-checker/default.htm

10 https://www.drugs.com/drug_interactions.html

11 https://go.drugbank.com/drug-interaction-checker

